# Pamrevlumab did not meet its primary endpoint for non-ambulatory patients with Duchenne Muscular Dystrophy: the LELANTOS-1 trial

**DOI:** 10.1101/2025.02.17.25321407

**Authors:** Anne M. Connolly, Brenda L. Wong, Han C. Phan, Eugenio Mercuri, Xiaotang Cai, Cuixia Tian, Silvana De Lucia, Stanley F. Nelson, Yann Péréon, Stefano C. Previtali, Steven Wang, Olga V. Gambetti, John F. Brandsema, Ewa Carrier, the LELANTOS-1 study investigators

**Author notes:** **Correspondence to:** Anne M. Connolly, MD, Nationwide Children’s Hospital, The Ohio State University College of Medicine, 700 Children’s Drive, Columbus, OH 43205 USA.

## Abstract

**Background:** In Duchenne muscular dystrophy (DMD), fibrosis is linked to connective tissue growth factor (CTGF) overexpression. Pamrevlumab, a fully human monoclonal antibody, inhibits CTGF activity and showed promise as a DMD treatment in a phase 2 trial.

**Objective:** LELANTOS-1 (NCT04371666) was a global phase 3 study of the safety and efficacy of pamrevlumab for non-ambulatory males ≥12 years old with DMD.

**Methods:** Patients were randomized 1:1 to pamrevlumab 35 mg/kg every 2 weeks for 52 weeks or placebo. All received a stable corticosteroid regimen (deflazacort or prednisone/prednisolone). Primary endpoint was total Performance of Upper Limb (PUL) v2.0 score change from baseline to Week 52 in the modified intent-to-treat (mITT) set (baseline PUL score ≥2). Treatment-emergent adverse events (TEAEs) were noted. Patients who completed the main study period were eligible to enroll in the open-label extension (OLE).

**Results:** Ninety-eight patients (mean [SD] age, 15.5 [2.57] years) enrolled; 48 received pamrevlumab and 49 received placebo. Between-group baseline characteristics were similar. In the mITT set (mean [SD] age, 15.5 [2.64] years; pamrevlumab, n=41; placebo, n=42), the total PUL v2.0 score change was not significantly different (*p*=0.8802). The pamrevlumab group had more grip strength deterioration than placebo in dominant (*p*=0.0161) and nondominant hands (*p*=0.0052). Most patients (pamrevlumab, n=45/48 [93.8%]; placebo, n=48/49 [98.0%]) experienced TEAEs (most mild/moderate). One death occurred in the pamrevlumab group (unrelated to study drug). The OLE mITT set included 72 patients. OLE efficacy and safety were consistent with the main study period. No deaths occurred during the OLE.

**Conclusions:** Pamrevlumab failed to meet the primary endpoint. Its future as a DMD treatment is uncertain.

**Trial Registration:** ClinicalTrials.gov Identifier: NCT04371666

## INTRODUCTION

In Duchenne muscular dystrophy (DMD), a loss-of-function mutation in the *DMD* gene leads to the absence of functional dystrophin, the protein that is essential for the strength, stability, and endurance of muscle fibers as they contract and relax [1–3]. The resulting lack of dystrophin leads to progressive muscle deterioration and loss of ambulation by adolescence. Patients with DMD are generally wheelchair bound before they develop significant respiratory muscle weakness [4–6]. Respiratory complications are the primary cause of morbidity and mortality in DMD as progressive respiratory muscle weakness leads to hypoventilation and/or recurrent atelectasis and pneumonia, secondary to decreased cough effectiveness [1, 7, 8].

Currently, there are no curative treatments for DMD. Corticosteroids are the standard of care and have been shown to delay loss of ambulation and improve respiratory function [4, 8–12]. Exon-skipping therapy is a promising approach available for patients with specific mutations in *DMD*.^3,13^ By modifying splicing and shifting the reading frame, a translatable transcript is created and dystrophin production can be partially restored [2, 3]. However, exon-skipping therapy requires a mutation-specific approach. Currently approved therapies target exon 51, exon 53, and exon 45, and mutations in these regions affect 14%, 10%, and 9% of patients with DMD [2, 3]. The first gene therapy for DMD was approved in 2023 for patients aged 4 to 5 years with a specific *DMD* mutation [14]. This therapy uses an adeno-associated virus vector to deliver a gene encoding a shorter-than-normal dystrophin protein. Overall, progress in DMD treatment has been promising, but many patients remain ineligible for newer, individualized treatments.

Pamrevlumab is a fully human monoclonal antibody that binds to and inhibits the activity of connective tissue growth factor (CTGF), which is believed to play a substantial role in fibrosis [15–18]. In the open-label phase 2 MISSION study of patients with DMD, pamrevlumab demonstrated slower disease progression than expected based on natural disease history and was well tolerated [19]. The results of the phase 2 study supported the initiation of this phase 3 study of the safety and efficacy of pamrevlumab in combination with systemic corticosteroids for non-ambulatory males with DMD.

The objective of the phase 3 LELANTOS-1 trial (NCT04371666) was to evaluate the safety and efficacy of pamrevlumab for the treatment of non-ambulatory males with DMD. Its companion trial, LELANTOS-2 (NCT04632940) [20], was designed to evaluate pamrevlumab for ambulatory males with DMD.

## METHODS

### Study design

LELANTOS-1 was a phase 3, randomized, double-blind, placebo-controlled, multicenter, multinational study. Patients were randomized 1:1 to receive pamrevlumab 35 mg/kg by intravenous (IV) administration or placebo every 2 weeks for 52 weeks (**Figure 1**). All patients were required to be on a stable dose of systemic corticosteroids (deflazacort or prednisone/prednisolone) before and during the study. Patients who completed the treatment period were eligible to enroll in an open-label extension (OLE) period, in which all patients received pamrevlumab IV 35 mg/kg every 2 weeks until the last patient from the phase 3 study completed the 52-week treatment period or until the OLE was terminated based on sponsor decision or commercial availability of pamrevlumab.

**Figure 1.**
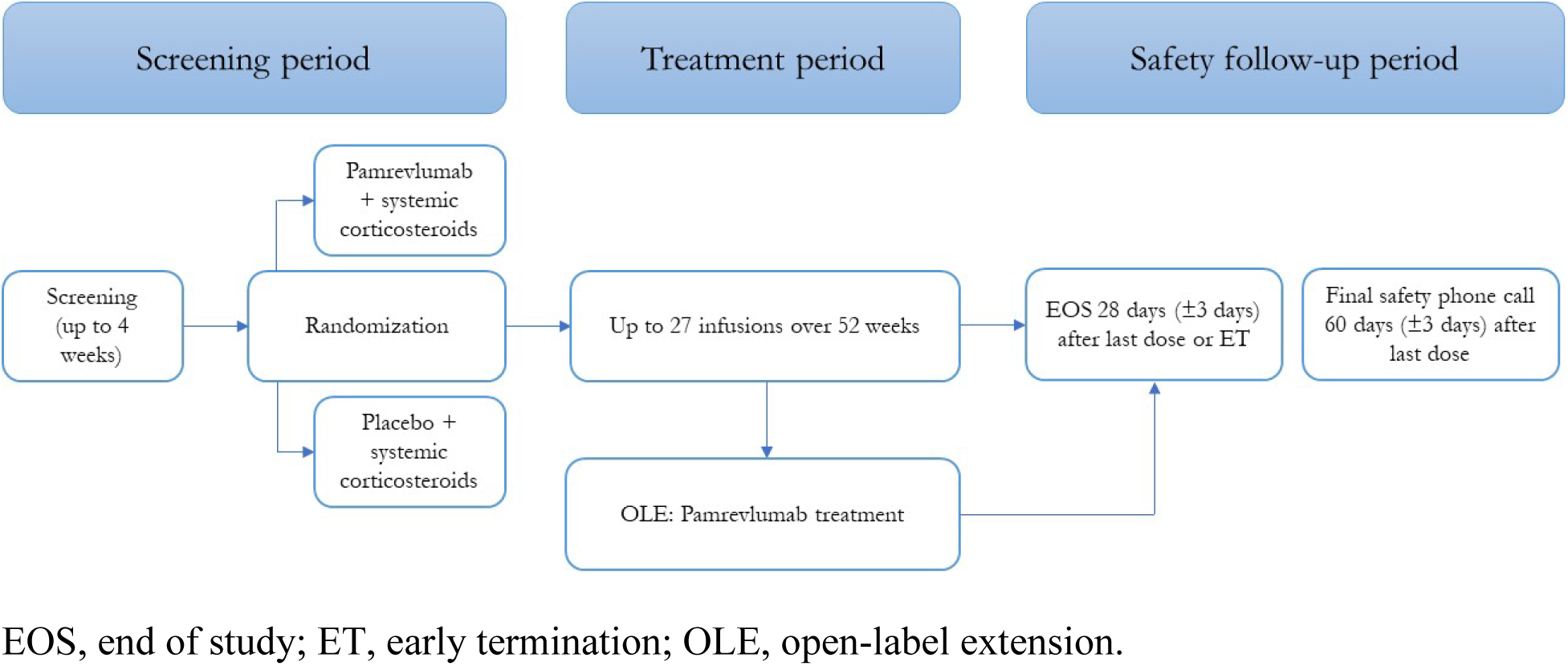
Study design.

### Patients

Males with DMD were included in the study if they were at least 12 years old and non-ambulatory at the time of screening. Patients were excluded if they had recent use of approved exon-skipping therapies for DMD. Full inclusion/exclusion criteria are listed in the **Supplement**.

Several analysis populations were considered. The Intent-to-Treat (ITT) set included all randomized patients. Patients were analyzed according to their randomized treatment group. The modified ITT (mITT) set (enriched for expected change) included patients from the ITT set with a performance of upper limb (PUL) entry score ≥2 (i.e., excluding PUL entry scores of 1) at baseline. The Per-Protocol set included all randomized patients who completed at least 24 doses of treatment who had a baseline and at least one post-baseline PUL assessment, did not discontinue early from treatment or study, and had no major protocol deviation(s) that significantly impacted efficacy analyses. The Safety Analysis Set included all patients who received any dose of study medication. All safety data were analyzed using the Safety Analysis Set. Patients were analyzed according to the treatment actually received.

### Endpoints

The primary efficacy endpoint was change in total score of PUL v2.0 from baseline to Week 52. Secondary efficacy endpoints were changes from baseline to Week 52 in ppFVC (assessed by spirometry), total score of mid-level (elbow) PUL, grip strength (assessed by handheld myometry), and left ventricular ejection fraction percentage (LVEF%; assessed by magnetic resonance imaging [MRI]). The mean observed z scores for each of three composite endpoints (PUL and ppFVC; PUL, ppFVC, and grip strength; PUL, ppFVC, grip strength, and LVEF%) were calculated.

Exploratory endpoints included changes from baseline to Week 52 in the subscores of regional dimensions (high-level [shoulder] and distal-level [wrist and hand]) of PUL, Duchenne Video Assessment severity percentage, percent-predicted forced expiratory volume in 1 second, percent-predicted peak expiratory flow assessed by spirometry, and progression of dilated cardiomyopathy assessed by MRI and grouped via genetic analysis at Week 52. MRI assessments of fibrosis included changes from baseline to Week 52 in fibrosis score of the biceps brachii, cardiac fibrosis score assessed by Late Gadolinium Enhancement, and myocardial circumferential strain (global circumferential strain) percentage assessed by cardiac MRI.

Safety endpoints included all treatment-emergent adverse events (TEAEs), treatment-emergent serious adverse events (TESAEs), clinically significant laboratory abnormalities, discontinuation of treatment due to TEAEs, hypersensitivity/anaphylactic reactions, and infusion reactions. Adverse events were coded using MedDRA version 26.0 or higher. The number (percentage) of patients with hospitalizations due to any serious adverse event (SAE) with a pulmonary or cardiac cause was reported, as well as the number (percentage) of patients with bone fractures. Ulna length measurements for indirect measure of growth velocity (cm/year) for patients under 18 years of age were also reported.

### Statistical analysis

All analyses were performed using SAS version 9.4 (Cary, NC, USA) or higher. Frequency distributions (number and percentage of patients) were presented for categorical variables. For continuous variables, descriptive statistics, including n, mean, standard deviation (SD), median, minimum, and maximum were presented. Unless otherwise stated, all confidence intervals (CIs) were two-sided 95% CIs. Analysis of all efficacy endpoints was based on the mITT set unless otherwise specified. Sensitivity analyses on the mITT set were also performed on primary and secondary endpoints.

### Ethics

Informed consent was obtained from each patient or their legal guardian, and the study was approved by the respective Institutional Review Boards of each participating study site (**Table S1**).

This study was conducted in accordance with the Declaration of Helsinki, Good Clinical Practice (GCP), the International Council for Harmonisation E6 Guidance for GCP, and any other applicable local health and regulatory requirements.

## RESULTS

### Patient disposition

One hundred fifty patients from 53 study sites were screened (**Figure 2**). Ninety-eight patients were enrolled and randomized, and 97 of these patients received at least one dose of a study treatment (pamrevlumab or placebo). Eighty-nine patients completed the treatment period and nine discontinued treatment early. Ninety patients completed the study and 86 entered the OLE. The analysis sets are presented in **Table 1**.

**Figure 2.**
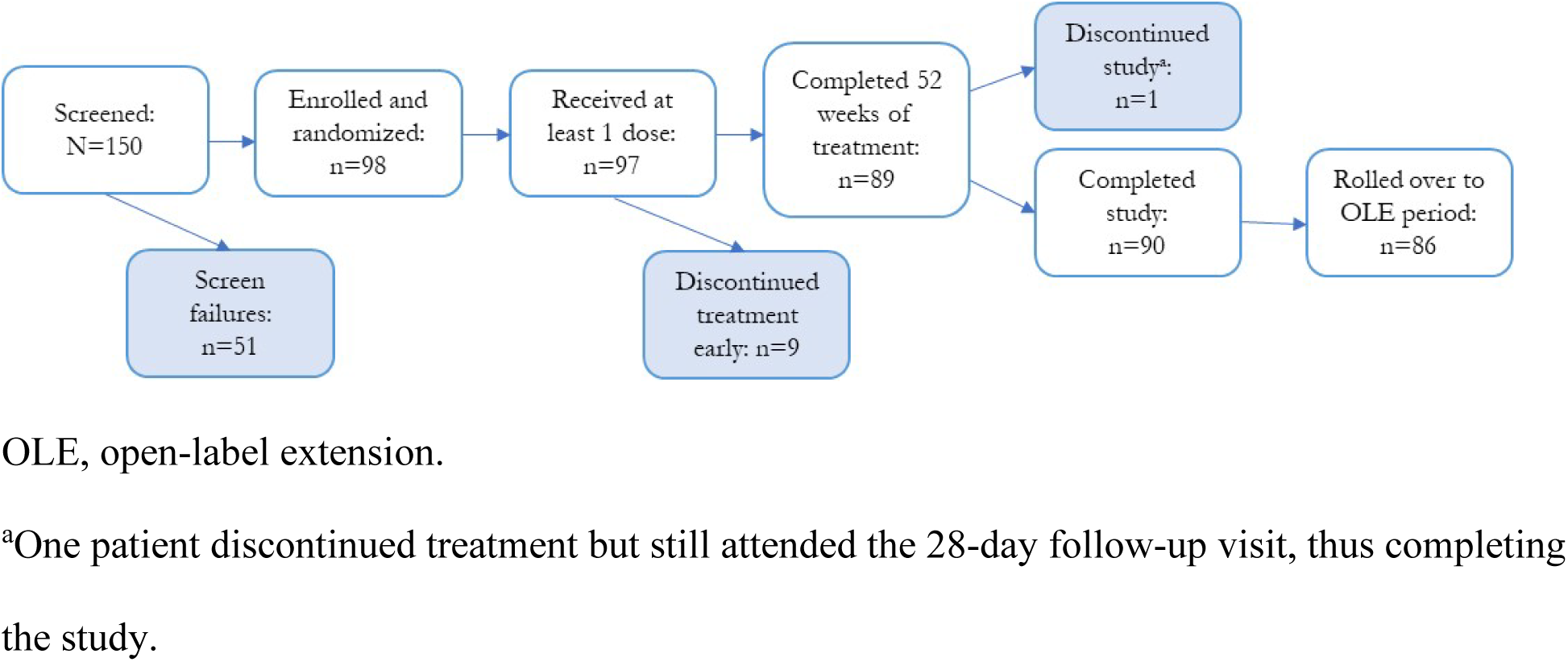
Patient disposition.

**Table 1.** Analysis sets.

| Populations | Main study period |  |  | OLE |  |  |
| --- | --- | --- | --- | --- | --- | --- |
|  | Pamrevlumab | Placebo | Overall | Pamrevlumab | Placebo | Overall |
| **ITT set, n (%)** | 49 (100) | 49 (100) | 98<br>(100) | 43 (100) | 43 (100) | 86<br>(100) |
| **mITT set, n (%)** | 41 (83.7) | 42<br>(85.7) | 83<br>(84.7) | 35 (81.4) | 37<br>(86.0) | 72<br>(83.7) |
| **Per-protocol set, n (%)** | 37 (75.5) | 45<br>(91.8) | 82<br>(83.7) | N/A | N/A | N/A |
| **Safety analysis set, n (%)** | 48 (98.0) | 49 (100) | 97<br>(99.0) | 40 (93.0) | 38<br>(88.4) | 78<br>(90.7) |

ITT, intent-to-treat; mITT, modified intent-to-treat; N/A, not applicable; OLE, open-label extension.

Demographics and disease characteristics were evenly balanced between the pamrevlumab and placebo groups (**Table 2**). The average age of the entire population was 15.5 years. Most patients were White (n=77; 78.6%) and not Hispanic or Latino (n=85; 86.7%). Treatment compliance was high in both groups (median [range]: pamrevlumab, 96.24% [14.8–100.1%]; placebo, 96.51% [47.3–100.6%]).

**Table 2.** Demographic, clinical, and disease characteristics.

| Variable |  | ITT set |  |  | mITT set |  |  |
| --- | --- | --- | --- | --- | --- | --- | --- |
|  |  | Pamrevlumab<br>(n=49) | Placebo<br>(n=49) | Overall<br>(N=98) | Pamrevlumab<br>(n=41) | Placebo<br>(n=42) | Overall<br>(N=83) |
| Age, years | Mean (SD) | 15.6 (2.74) | 15.5<br>(2.42) | 15.5<br>(2.57) | 15.6 (2.92) | 15.5 (2.38) | 15.5 (2.64) |
|  | Median (range) | 15.0 (12–27) | 15.0 (12–<br>20) | 15.0 (12–<br>27) | 15.0 (12–27) | 15.0 (12–<br>20) | 15.0 (12–27) |
| Age group, n<br>(%) | ≤16 years old | 37 (75.5) | 34 (69.4) | 71 (72.4) | 31 (75.6) | 29 (69.0) | 60 (72.3) |
|  | >16 years old | 12 (24.5) | 15 (30.6) | 27 (27.6) | 10 (24.4) | 13 (31.0) | 23 (27.7) |
| Sex, n (%) | Male | 49 (100) | 49 (100) | 98 (100) | 41 (100) | 42 (100) | 83 (100) |
| Ethnicity, n<br>(%) | Hispanic or Latino | 2 (4.1) | 4 (8.2) | 6 (6.1) | 1 (2.4) | 4 (9.5) | 5 (6.0) |
|  | Not Hispanic or Latino | 44 (89.8) | 41 (83.7) | 85 (86.7) | 37 (90.2) | 34 (81.0) | 71 (85.5) |
|  | Not reported | 3 (6.1) | 4 (8.2) | 7 (7.1) | 3 (7.3) | 4 (9.5) | 7 (8.4) |
| Race, n (%) | American Indian or<br>Alaska Native | 0 | 0 | 0 | 0 | 0 | 0 |
|  | Asian | 6 (12.2) | 9 (18.4) | 15 (15.3) | 5 (12.2) | 6 (14.3) | 11 (13.3) |
|  | Black or African<br>American | 0 | 2 (4.1) | 2 (2.0) | 0 | 2 (4.8) | 2 (2.4) |
|  | Native Hawaiian or<br>other Pacific Islander | 0 | 0 | 0 | 0 | 0 | 0 |
|  | White | 42 (85.7) | 35 (71.4) | 77 (78.6) | 35 (85.4) | 31 (73.8) | 66 (79.5) |
|  | Other | 1 (2.0) | 3 (6.1) | 4 (4.1) | 1 (2.4) | 3 (7.1) | 4 (4.8) |
| BMI, kg/m<sup>2</sup> | Mean (SD) | 25.27 (5.859) | 25.20<br>(6.238) | 25.24<br>(6.020) | 25.44 (6.200) | 24.85<br>(5.768) | 25.14<br>(5.956) |
|  |  | Pamrevlumab<br>(n=49) | Placebo<br>(n=49) | Overall<br>(N=98) | Pamrevlumab<br>(n=41) | Placebo<br>(n=42) | Overall<br>(N=83) |
|  | Median (range) | 25.59 (16.1–<br>37.4) | 25.14<br>(12.2–<br>39.1) | 25.45<br>(12.2–<br>39.1) | 25.65 (16.1–<br>37.4) | 24.66<br>(12.2–<br>39.1) | 25.43 (12.2–<br>39.1) |
| Dominant arm,<br>n (%) | Left | 9 (18.4) | 6 (12.2) | 15 (15.3) | 7 (17.1) | 6 (14.3) | 13 (15.7) |
|  | Right | 40 (81.6) | 43 (87.8) | 83 (84.7) | 34 (82.9) | 36 (85.7) | 70 (84.3) |
| *Disease history and characteristics* |  |  |  |  |  |  |  |
| Age at DMD<br>diagnosis | Mean (SD) | 4.8 (2.58) | 5.3 (2.58) | 5.0 (2.58) | 4.9 (2.57) | 5.5 (2.42) | 5.2 (2.50) |
|  | Median (range) | 5.0 (0–11) | 6.0 (1–11) | 5.0 (0–11) | 5.0 (1–11) | 6.0 (2–11) | 5.0 (1–11) |
| Age at loss of<br>ambulation | Mean (SD) | 12.3 (2.39) | 11.8<br>(2.70) | 12.1<br>(2.55) | 12.5 (2.39) | 11.8 (2.84) | 12.2 (2.63) |
|  | Median (range) | 12.0 (8–20) | 12.0 (3–<br>17) | 12.0 (3–<br>20) | 12.0 (8–20) | 11.0 (3–<br>17) | 12.0 (3–20) |
| Years since<br>loss of<br>ambulation | Mean (SD) | 3.2 (2.02) | 3.6 (2.12) | 3.4 (2.07) | 3.0 (2.09) | 3.6 (2.22) | 3.3 (2.16) |
|  | Median (range) | 3.0 (0–11) | 3.0 (0–10) | 3.0 (0–11) | 3.0 (0–11) | 3.0 (0–10) | 3.0 (0–11) |
| Brooke score | Mean (SD) | 3.0 (1.07) | 3.1 (1.12) | 3.0 (1.09) | 2.7 (0.85) | 2.9 (1.03) | 2.8 (0.95) |
|  | Median (range) | 3.0 (1–5) | 3.0 (1–5) | 3.0 (1–5) | 3.0 (1–5) | 3.0 (1–5) | 3.0 (1–5) |
| ppFVC | Mean (SD) | 65.90 (11.138) | 66.57<br>(11.300) | 66.24<br>(11.166) | 67.39 (10.900) | 67.58<br>(11.482) | 67.49<br>(11.130) |
|  | Median (range) | 65.80 (41–<br>82.6) | 67.50<br>(42.4–<br>88.2) | 66.60<br>(41–88.2) | 69.00 (41–<br>82.6) | 68.35<br>(42.4–<br>88.2) | 68.50 (41–<br>88.2) |
| Corticosteroid<br>use, n (%) | Prednisone/prednisolone | 14 (28.6) | 15 (30.6) | 29 (29.6) | 11 (26.8) | 11 (26.2) | 22 (26.5) |
|  | Deflazacort | 35 (71.4) | 34 (69.4) | 69 (70.4) | 30 (73.2) | 31 (73.8) | 61 (73.5) |
|  |  | Pamrevlumab<br>(n=49) | Placebo<br>(n=49) | Overall<br>(N=98) | Pamrevlumab<br>(n=41) | Placebo<br>(n=42) | Overall<br>(N=83) |
|  | Daily regimen | 41 (83.7) | 43 (87.8) | 84 (85.7) | 35 (85.4) | 36 (85.7) | 71 (85.5) |
|  | Other regimen | 8 (16.3) | 6 (12.2) | 14 (14.3) | 6 (14.6) | 6 (14.3) | 12 (14.5) |
| Age at first use<br>of<br>corticosteroids,<br>years | Mean (SD) | 6.6 (2.28) | 6.8 (2.50) | 6.7 (2.38) | 6.8 (2.39) | 6.7 (2.22) | 6.7 (2.29) |
|  | Median (range) | 6.0 (2–12) | 7.0 (2–14) | 6.5 (2–14) | 7.0 (2–12) | 7.0 (2–13) | 7.0 (2–13) |
| Years since<br>first use of<br>corticosteroids | Mean (SD) | 9.0 (3.34) | 8.7 (3.54) | 8.8 (3.43) | 8.8 (3.49) | 8.8 (3.39) | 8.8 (3.42) |
|  | Median (range) | 9.0 (3–18) | 8.0 (2–17) | 9.0 (2–18) | 8.0 (3–18) | 8.5 (2–17) | 8.0 (2–18) |
| LVEF% |  | *n=40* | *n=45* | *n=85* | *n=33* | *n=38* | *n=71* |
|  | Mean (SD) | 55.30 (6.349) | 52.87<br>(6.428) | 54.01<br>(6.469) | 54.83 (6.781) | 53.01<br>(6.719) | 53.86<br>(6.762) |
|  | Median (range) | 55.66 (39.1–<br>66.9) | 53.72<br>(38.3–<br>63.3) | 54.50<br>(38.3–<br>66.9) | 54.81 (39.1–<br>66.9) | 54.12<br>(38.3–<br>63.3) | 54.50 (38.3–<br>66.9) |
| PUL v2.0 entry<br>score at<br>baseline, n (%) | 1 | 8 (16.3) | 7 (14.3) | 15 (15.3) | 0 | 0 | 0 |
|  | 2 | 3 (6.1) | 5 (10.2) | 8 (8.2) | 3 (7.3) | 5 (11.9) | 8 (9.6) |
|  | 3 | 17 (34.7) | 20 (40.8) | 37 (37.8) | 17 (41.5) | 20 (47.6) | 37 (44.6) |
|  | 4 | 7 (14.3) | 7 (14.3) | 14 (14.3) | 7 (17.1) | 7 (16.7) | 14 (16.9) |
|  | 5 | 9 (18.4) | 4 (8.2) | 13 (13.3) | 9 (22.0) | 4 (9.5) | 13 (15.7) |
|  | 6 | 5 (10.2) | 6 (12.2) | 11 (11.2) | 5 (12.2) | 6 (14.3) | 11 (13.3) |
| PUL v2.0 total<br>score | Mean (SD) | 25.8 (8.20) | 24.8<br>(7.42) | 25.3<br>(7.80) | 27.7 (7.60) | 26.0 (7.23) | 26.8 (7.42) |
|  | Median (range) | 25.0 (13–42) | 23.0 (14–<br>42) | 23.5 (13–<br>42) | 27.0 (16–42) | 24.0 (14–<br>42) | 26.0 (14–42) |
|  |  | Pamrevlumab<br>(n=49) | Placebo<br>(n=49) | Overall<br>(N=98) | Pamrevlumab<br>(n=41) | Placebo<br>(n=42) | Overall<br>(N=83) |
| PUL v2.0,<br>high-level<br>score | Mean (SD) | *n*=38<br>4.4 (3.77) | *n*=37<br>3.6 (3.80) | *n*=75<br>4.0 (3.78) | *n*=38<br>4.4 (3.77) | *n*=37<br>3.6 (3.80) | *n*=75<br>4.0 (3.78) |
|  | Median (range) | 4.0 (0–12) | 2.0 (0–12) | 3.0 (0–12) | 4.0 (0–12) | 2.0 (0–12) | 3.0 (0–12) |
| PUL v2.0,<br>mid-level score | Mean (SD) | 11.3 (3.88) | 10.7<br>(3.60) | 11.0<br>(3.73) | 12.2 (3.50) | 11.5 (3.31) | 11.8 (3.41) |
|  | Median (range) | 11.0 (4–17) | 10.0 (3–<br>17) | 10.5 (3–<br>17) | 12.0 (6–17) | 11.0 (3–<br>17) | 11.0 (3–17) |
| PUL v2.0,<br>distal-level<br>score | Mean (SD) | 11.1 (1.29) | 11.3<br>(1.03) | 11.2<br>(1.17) | 11.4 (1.16) | 11.4 (1.01) | 11.4 (1.08) |
|  | Median (range) | 11.0 (8–13) | 11.0 (9–<br>13) | 11.0 (8–<br>13) | 11.0 (8–13) | 11.0 (9–<br>13) | 11.0 (8–13) |

BMI, body mass index; DMD, Duchenne muscular dystrophy; ITT, intent-to-treat; LVEF%, left ventricular ejection fraction
percentage; mITT, modified intention-to-treat; ppFVC, percent-predicted forced vital capacity; PUL, performance of upper limb; SD, standard deviation.

### Efficacy

The difference in change from baseline in total PUL v2.0 score was not different between the treatment groups (**Figure 3**). The mean (SD) change in total PUL v2.0 score was –2.036 (0.4471) for the pamrevlumab group versus –2.119 (0.3367) for the placebo group; the least-squares (LS) mean difference was 0.083 (*p*=0.8802) (**Table 3**). Therefore, the primary endpoint was not met. Sensitivity analyses revealed similar results, and subgroup analyses of the primary endpoint did not demonstrate significant differences between treatment groups. Of note, for the primary endpoint, patients with a baseline PUL total score higher than the median (23.5) responded more favorably to pamrevlumab (LS mean [95% CI] change: pamrevlumab, –1.361 [– 2.556, –0.166]; placebo, –2.822 [–4.054, –1.590]), and the difference approached statistical significance (*p*=0.08); conversely, patients with baseline PUL total scores equal to or below the median showed less deterioration on placebo (LS mean [95% CI] change: pamrevlumab, –2.499 [–3.964, –1.034]; placebo, –1.345 [–2.087, –0.602]; *p*=0.14).

**Figure 3.**
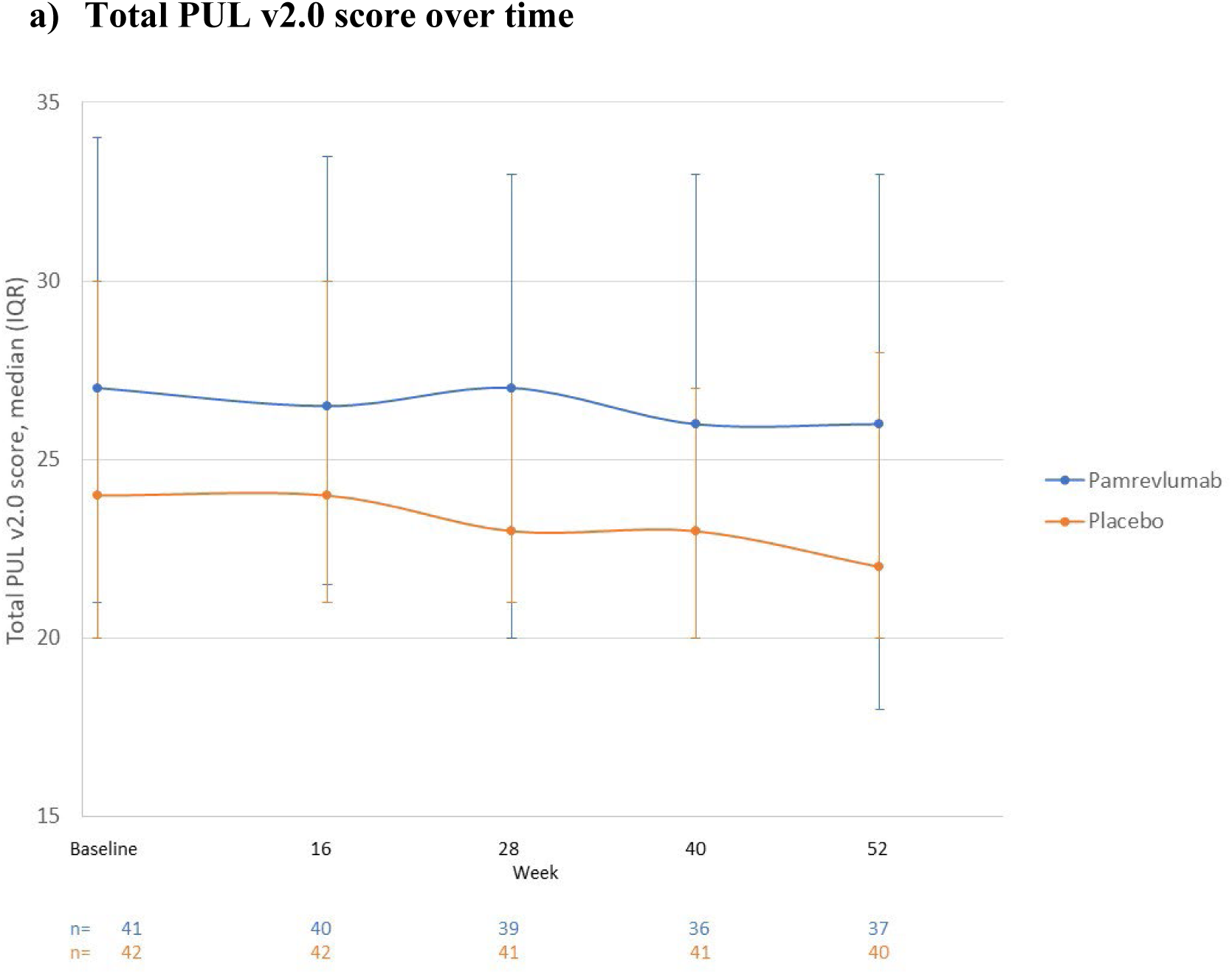
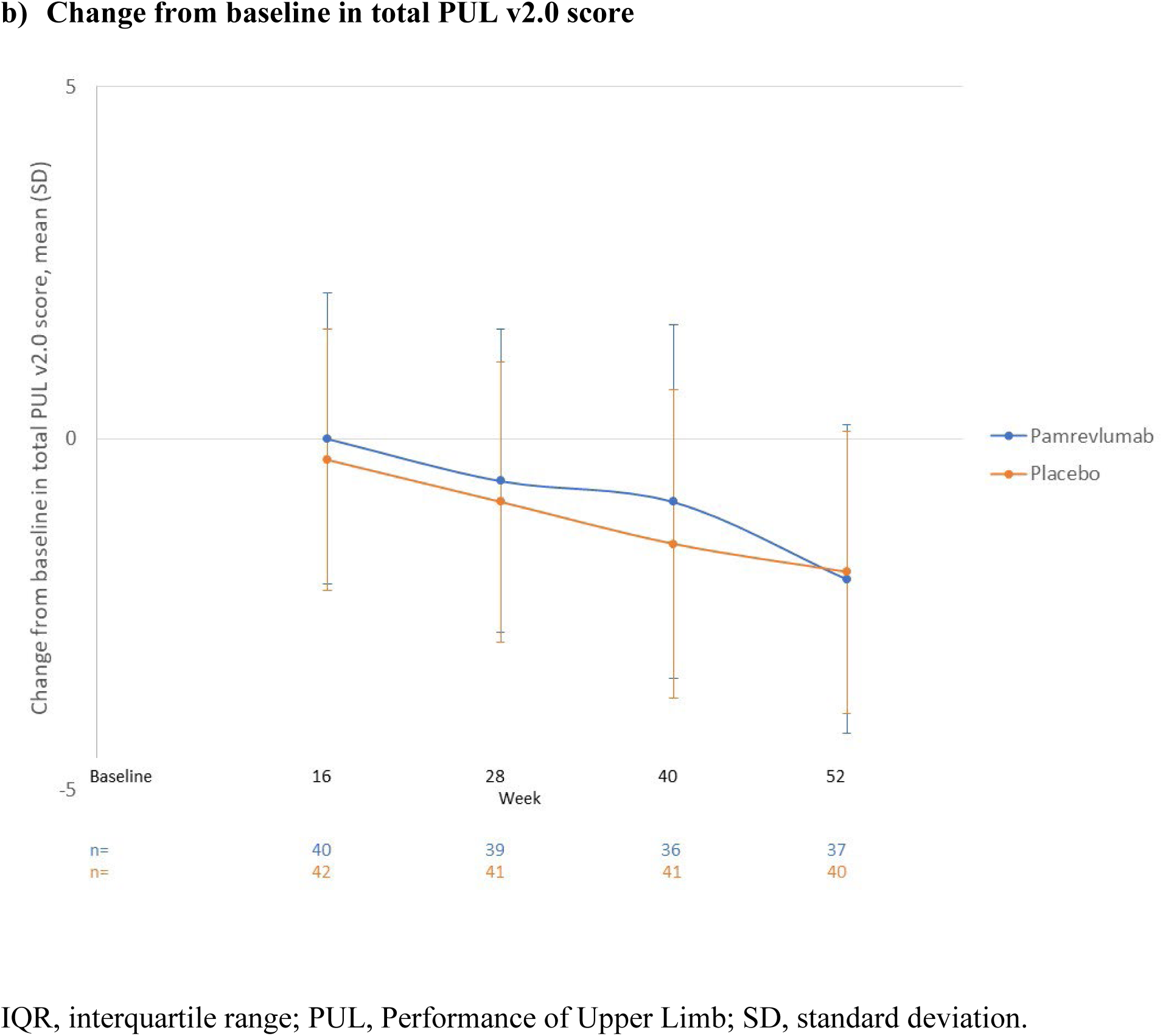
Total PUL v2.0 score during the main study period.

**Table 3.** Primary and secondary efficacy endpoints for mITT population in main study period and OLE.

| Parameter | Change from baseline at Week 52,<br>LS mean (SE) |  |  | Change from baseline at OLE<br>Week 48, LS mean (SE) |  |  |
| --- | --- | --- | --- | --- | --- | --- |
|  | Pamrevlumab<br>(n=41) | Placebo<br>(n=42) | p-<br>value | Pamrevlumab<br>(n=35) | Placebo<br>(n=37) | p-<br>value |
| **PUL v2.0**<br>**total score** | n=38<br>-2.036 (0.4471) | n=40<br>-2.119<br>(0.3367) | 0.8802 | n=13<br>-3.871 (0.6400) | n=13<br>-3.794<br>(0.5939) | 0.9298 |
| **ppFVC** | n=38<br>-8.349 (1.5760) | n=40<br>-5.989<br>(1.2233) | 0.2404 | n=13<br>-13.520<br>(1.7764) | n=13<br>-10.483<br>(1.6877) | 0.2165 |
| **Mid-level of**<br>**PUL v2.0**<br>**total score** | n=38<br>-1.021 (0.3261) | n=40<br>-1.200<br>(0.2651) | 0.6675 | n=13<br>-1.972 (0.4235) | n=13<br>-1.906<br>(4.051) | 0.9104 |
| **Grip strength**<br>**by dominant**<br>**hand, N** | n=37<br>-7.570 (2.0989) | n=39<br>-0.072<br>(2.2065) | 0.0161 | n=12<br>-8.364 (4.0941) | n=10<br>-5.191<br>(4.0898) | 0.5843 |
| **Grip strength**<br>**by non-**<br>**dominant**<br>**hand, N** | n=36<br>-7.627 (1.8893) | n=39<br>-0.012<br>(1.8546) | 0.0052 | n=12<br>-7.164 (4.2512) | n=10<br>-8.269<br>(4.1007) | 0.8517 |
| **LVEF%** | n=33<br>-0.499 (0.9750) | n=37<br>-1.114<br>(0.9204) | 0.6490 | n=8<sup>a</sup><br>-0.489 (2.0822) | n=10<sup>a</sup><br>-2.403<br>(1.7270) | 0.4889 |

LS, least-squares; LVEF%, left ventricular ejection fraction percentage; mITT, modified intent-
to-treat; OLE, open-label extension; PUL, performance of upper limb; SE, standard error.
<sup>a</sup>Last available data: OLE Week 52.

No clinically meaningful between-group findings in the secondary endpoints were observed for change in ppFVC, mid-level of PUL score, or LVEF% (**Table 3**). For the secondary endpoint of grip strength, the pamrevlumab group experienced more deterioration than the placebo group in both hands, and the difference was statistically significant (dominant hand, *p*=0.0161; nondominant hand, *p*=0.0052) (**Table 3**). Ad-hoc analyses using subgroups with PUL total scores above and below the median were also performed on the secondary endpoints, with mixed results. For the change in mid-level of the PUL, pamrevlumab patients had more favorable results compared with placebo than in the full mITT set (LS mean [95% CI] difference, 1.6 [0.04, 3.1]; *p*=0.045). The changes in ppFVC, grip strength, and LVEF% were consistent with their respective mITT set results.

None of the three composites of endpoints showed statistical significance between the pamrevlumab group and the placebo group. There were no noteworthy trends among secondary endpoint subgroup analyses. Results aligned with either the full population’s secondary endpoint analyses and/or the primary endpoint subgroup findings.

None of the exploratory endpoint analyses demonstrated statistically significant differences between treatment groups. OLE efficacy results were consistent with results from the main study, and no strong trends were detected (**Table 3**).

### Safety

The incidence of TEAEs was similar in each treatment group; 45 patients (93.8%) in the pamrevlumab group and 48 (98.0%) in the placebo group experienced at least one TEAE during the study period (**Table 4**). The most common TEAEs for the pamrevlumab treatment group with a ≥5% greater incidence than the placebo group were headache (52% vs 20%), pyrexia (25% vs 8%), vomiting (21% vs 10%), nausea (17% vs 6%), and back/flank pain (17% vs 6%). More patients in the pamrevlumab treatment group had TEAEs considered related to the study drug (44% vs 25%) (**Table 5**). Nine patients (18.8%) in the pamrevlumab group and five (10.2%) in the placebo group experienced Grade ≥3 TEAEs (**Table 4**). Two TEAEs Grade ≥3 were considered related to pamrevlumab, one Grade 3 TEAE of troponin I increase and one Grade 4 TEAE of drug hypersensitivity.

**Table 4.** Safety summary.

| Category | Main study period |  | OLE period |  |
| --- | --- | --- | --- | --- |
|  | Pamrevlumab<br>(n=48), n (%) | Placebo<br>(n=49), n (%) | Pamrevlumab<br>(n=43), n (%) | Placebo<br>(n=43), n (%) |
| Any TEAE | 45 (93.8) | 48 (98.0) | 33 (76.7) | 34 (79.1) |
| Any TEAE leading to discontinuation of study medication | 3 (6.3) | 1 (2.0) | 0 | 0 |
| Any TEAE leading to interruption of study medication | 23 (47.9) | 15 (30.6) | 7 (16.3) | 4 (9.3) |
| Any TEAE related to study medication (as determined by the investigator) | 21 (43.8) | 12 (24.5) | 4 (9.3) | 10 (23.3) |
| TEAE by maximum severity |  |  |  |  |
| Grade 1/mild | 17 (35.4) | 21 (42.9) | 17 (39.5) | 13 (30.2) |
| Grade 2/moderate | 19 (39.6) | 22 (44.9) | 14 (32.6) | 16 (37.2) |
| Grade 3/severe | 5 (10.4) | 5 (10.2) | 2 (4.7) | 5 (11.6) |
| Grade 4/life threatening | 3 (6.3) | 0 | 0 | 0 |
| Grade 5/death | 1 (2.1) | 0 | 0 | 0 |
| Any TESAE | 8 (16.7) | 5 (10.2) | 3 (7.0) | 2 (4.7) |
| Any TESAE related to study medication (as determined by the investigator) | 1 (2.1) | 2 (4.1) | 0 | 0 |
| Any TESAE leading to death | 1 (2.1) | 0 | 0 | 0 |
| All reported deaths | 1 (2.1) | 0 | 0 | 0 |

OLE, open-label extension; TEAE, treatment-emergent adverse event; TESAE, treatment-emergent serious adverse event.

**Table 5.** TEAEs with an incidence of ≥5% in either treatment group during main study period.

| **System Organ Class**<br>**Preferred Term** | **Pamrevlumab**<br>**(n=48)**<br>**n (%)** | **Placebo**<br>**(n=49)**<br>**n (%)** |
| --- | --- | --- |
| Patients with any TEAEs with incidence $\geq 5\%$ in either treatment group | 42 (87.5) | 45 (91.8) |
| Cardiac disorders | 1 (2.1) | 5 (10.2) |
| Cardiomyopathy | 1 (2.1) | 5 (10.2) |
| Eye disorders | 3 (6.3) | 0 |
| Cataract | 3 (6.3) | 0 |
| Gastrointestinal disorders | 16 (33.3) | 11 (22.4) |
| Vomiting | 10 (20.8) | 5 (10.2) |
| Nausea | 8 (16.7) | 3 (6.1) |
| Diarrhea | 5 (10.4) | 4 (8.2) |
| General disorders and administration site conditions | 14 (29.2) | 4 (8.2) |
| Pyrexia | 12 (25.0) | 4 (8.2) |
| Pain | 4 (8.3) | 0 |
| Infections and infestations | 24 (50.0) | 28 (57.1) |
| COVID-19 | 19 (39.6) | 20 (40.8) |
| Nasopharyngitis | 3 (6.3) | 7 (14.3) |
| Upper respiratory tract infection | 6 (12.5) | 3 (6.1) |
| Rhinitis | 1 (2.1) | 4 (8.2) |
| Injury, poisoning, and procedural complications | 10 (20.8) | 8 (16.3) |
| Vascular access site bruising | 3 (6.3) | 4 (8.2) |
| Contusion | 4 (8.3) | 2 (4.1) |
| Femur fracture | 1 (2.1) | 3 (6.1) |
| Limb injury | 4 (8.3) | 0 |
| Ligament sprain | 0 | 3 (6.1) |
| Investigations | 3 (6.3) | 0 |
| Heart rate increased | 3 (6.3) | 0 |
| Musculoskeletal and connective tissue disorders | 9 (18.8) | 9 (18.4) |
| Back pain | 7 (14.6) | 3 (6.1) |
| Arthralgia | 2 (4.2) | 4 (8.2) |
| Pain in extremity | 2 (4.2) | 3 (6.1) |
| Nervous system disorders | 25 (52.1) | 11 (22.4) |
| Headache | 25 (52.1) | 10 (20.4) |
| Dizziness | 4 (8.3) | 2 (4.1) |
| Respiratory, thoracic, and mediastinal disorders | 6 (12.5) | 10 (20.4) |
| Cough | 2 (4.2) | 4 (8.2) |
| Oropharyngeal pain | 3 (6.3) | 3 (6.1) |
| Rhinorrhea | 1 (2.1) | 3 (6.1) |
| Skin and subcutaneous tissue disorders | 4 (8.3) | 5 (10.2) |
| Rash | 4 (8.3) | 5 (10.2) |
| Vascular disorders | 3 (6.3) | 2 (4.1) |
| Flushing | 3 (6.3) | 2 (4.1) |

TEAE, treatment-emergent adverse event.

The incidence of TESAEs was slightly greater in the pamrevlumab group than the placebo group (16.7% vs 10.2%). TESAEs largely coincided with Grade ≥3 TEAEs. Four TESAEs occurred in more than one patient: myocarditis (one in each group), pneumonia (one in each group), and femur fractures (one in pamrevlumab group, two in placebo group). One patient in the pamrevlumab group and two patients in the placebo group had TESAEs considered related to the study drug (pamrevlumab: drug hypersensitivity; placebo: acute respiratory failure and pneumonia in a single patient and myocarditis in another patient). One patient experienced a TESAE (cardiac arrest) that resulted in death. The patient, who was randomized to the pamrevlumab group, died 206 days after his first dose of pamrevlumab. Death was the sequelae of a cardiac arrest, in the context of pneumonia and a complicated clinical course in hospital. The event was considered not related to the study drug. There were no observed trends in TEAE or TESAE subgroups during the main study period.

There were no trends in hematology, chemistry, physical examinations, or vital signs results during the study. Electrocardiogram abnormalities were infrequent. Six patients in each group (pamrevlumab, 12.4%; placebo, 12.2%) experienced bone fractures. Annualized growth velocity based on ulna length measurements was similar between groups (mean [SD]: pamrevlumab, 2.95 [3.099] cm/year; placebo, 2.36 [2.160] cm/year).

No notable safety trends were observed in the OLE period (**Table 4**). TEAE incidence in the OLE period was approximately equal with the main study treatment groups (pamrevlumab, 77%; placebo, 79%). In the OLE period, two patients (4.7%) in the pamrevlumab treatment group and five patients (11.6%) in the placebo group had Grade 3 TEAEs. None were related to pamrevlumab. There were no Grade 4 or Grade 5 TEAEs. One patient in the pamrevlumab treatment group was hospitalized with a pulmonary and/or cardiac cause in the OLE period (Grade 2 dyspnea, a sequela of Grade 2 pneumonia, neither of which was considered related to study drug). There were no trends in hematology, chemistry, physical examinations, vital signs, or bone fractures, and no deaths occurred in the OLE period. Overall, the safety data were in line with expectations and did not reveal any new safety concerns. Pamrevlumab was generally safe and well tolerated.

## DISCUSSION

LELANTOS-1 did not meet its primary endpoint of change in total PUL v2.0 score during the main study period, and this trend continued through the OLE period. Sensitivity analyses using the ITT population and varying methodology for handling missing data demonstrated similar results. Subgroups had very small populations, limiting any conclusions on the basis of these analyses. Similarly, none of the secondary composite endpoints nor the exploratory endpoints demonstrated statistically significant differences between treatment groups. The results of the subgroup analyses of primary and secondary endpoints according to PUL above and below the median suggest that patients with less overall deterioration because of DMD may respond better to pamrevlumab than to placebo. However, the clinical significance of the results remains unclear due to small sample size, and this finding contradicts those of the companion LELANTOS-2 study, which indicated a subgroup of patients with less overall deterioration responding less favorably to pamrevlumab than placebo [20].

The results of the grip strength secondary endpoint were unexpected, and causation was unclear. Additionally, the average grip strength of the placebo group did not decline according to expectations based on the natural history of the disease. A one-year follow-up study in non-ambulatory patients with DMD showed a mean decline in grip strength of –2.7 N in the dominant hand and –3.0 N in the nondominant hand [21]. In LELANTOS-1 in the pamrevlumab group, the mean decline in grip strength was –7.50 N in the dominant hand and –7.627 N in the nondominant hand, and, in the placebo group, the changes were –0.0072 N and –0.0012 N, respectively, indicating that neither group had changes that aligned with expectations. Moreover, results of the pamrevlumab treatment group were in marked contrast with other endpoints as well as the results of the phase 2 open-label pamrevlumab study [19]. In the MISSION study, 21 non-ambulatory DMD patients had an improvement in grip strength of 1.0 N (95% CI: 5.9, 8.0) in the dominant hand and 1.9 N (95% CI: 4.9, 8.6) in the nondominant hand after one year of pamrevlumab treatment. In LELANTOS-1, interpatient variability in grip strength results was widely divergent. Of note, there were two outliers who may have had a strong effect on results. One patient discontinued the study due to a TEAE that resulted in death; results for Weeks 40 and 52 were therefore imputed to –60 N, which was much lower than any other patient’s results. Conversely, one patient in the placebo group had grip strength results that were much greater than all other patients; at Week 52, this patient’s results were >1.5 times the next best set of results, and three or four times the best results of most patients. Given these factors, the grip strength results are not considered an accurate representation of pamrevlumab’s clinical effect.

The primary endpoint of change in total score of PUL v2.0 from baseline to Week 52 was chosen for its validated utility in measuring upper limb motor performance across the wide spectrum of DMD severity. By enrolling non-ambulatory patients aged ≥12 years, this study focused on pamrevlumab’s efficacy in patients with DMD whose disease had advanced to loss of ambulation. Secondary and exploratory endpoints were chosen both to support primary efficacy findings and to examine the impact of pamrevlumab on multiple aspects of each body system affected by DMD (musculoskeletal, pulmonary, and cardiac).

No new safety concerns were identified and pamrevlumab was generally safe and well-tolerated in the study. The overall safety profile for TEAEs was consistent with that expected from a non-ambulatory DMD population and the safety profile of pamrevlumab.

Although pamrevlumab has a low safety risk to patients, its future as a DMD treatment remains uncertain. While certain subgroups had a better overall response to pamrevlumab, especially in regard to upper limb function, the results fell short of statistical significance in all but a few subgroups with very small sample sizes.

This study had several limitations. First, analyses of the primary endpoint included patients with disease that had progressed to loss of ambulation. Further, the subgroup analyses contained small numbers of patients, which limited interpretation and generalization of the findings. Finally, changes in grip strength, ppFVC, and LVEF% may need a much longer period of evaluation to establish significant differences.

## Acknowledgements

The authors would like to express their gratitude to the patients and their families for participating in this study. Medical writing support was provided by Jennifer L. Gibson, PharmD, of Kay Square Scientific (Newtown Square, PA, USA). This support was fully funded by FibroGen.

## Ethical Compliance

The study was conducted in accordance with the Declaration of Helsinki, Good Clinical Practice (GCP), the International Council for Harmonisation E6 Guidance for GCP, and any other applicable local health and regulatory requirements. The study was approved by the respective institutional review boards at each participating study site, and written informed consent was obtained from each patient or their legal guardian.

## Declaration of Funding

Funding for this study was provided by FibroGen, Inc. (San Francisco, CA, USA).

## Conflicts of Interest

**AMC** has received grants and reports serving on a advisory board for Edgewise Therapeutics and Sarepta. **BLW**, **YP**, **SW**, and **XC** report nothing to disclose. **HCP** served as a principal investigator for Applied Therapeutics, Avidity, Biogen, Biohaven, Dyne, Edgewise, Italfarmaco, NMD Pharma, NS Pharma, PepGen, Sarepta, Stealth, Takeda, Ultragenyx, and Zynerba. She also serves on the advisory board for Tyra and an advisory committee for the FDA. **EM** has received honoraria for scientific advisory boards and educational speaker fees from Biogen, Novartis, F. Hoffmann-La Roche, Sarepta Therapeutics, Santhera, PTC Therapeutics, Pfizer, Scholar Rock, and Cytokinetics, and has received research support grants from Biogen. **CT** received grants for serving as a site investigator for this study and for clinical trials and studies sponsored by Novartis, BioHaven, Capricor, Catabasis, MDA, PTC Therapeutics, and Sarepta, and received personal fees for serving as a member of advisory boards for Pfizer, Catalyst, ITF, and Sarepta, and grants for serving as co-PI for the National Institutes of Health–sponsored TSC-STEPS study. **SDL** has served as a PI for FibroGen and Genethon, on an advisory board for Solid, and as a consultant for Effik/Italfarmaco Group. **SFN** reports study fees paid to UCLA by FibroGen for this clinical trial. **SCP** hasreceived support from FibroGen for this clinical trial; support from Wave, Dyne, Argenx, and Jannsen for clinical trials; personal fees for scientific boards from Wave, Esperare, and Argenx; and grants from Fondazione ricerca regione lombardia for a clinical trial, and Telethon Italy for a natural history study. **OVG** was an employee of FibroGen at the time of the study, with restricted stock grants. **EC** is an employee of FibroGen and owns stock options. **JFB** received personal fees from AveXis/Novartis, Biogen, Dyne, Edgewise, FibroGen, Genentech, Momenta/Janssen, NS Pharma, Pfizer, PTC Therapeutics, Sarepta, Scholar Rock, Takeda, and WaVe. He has received grants from Alexion, Astellas, AveXis/Novartis, Biogen, Biohaven, Catabasis, CSL Behring, Cytokinetics, Dyne, Fibrogen, Genentech, ML Bio, Pfizer, PTC Therapeutics, Sarepta, and Scholar Rock.

## Data Availability Statement

FibroGen, Inc., is committed to data sharing and to furthering medical research and patient care. Based on scientific merit, requests from qualified external researchers for anonymized patient-level and study-level clinical trial data (including redacted clinical study reports) for medicines and indications approved in the United States and Europe will be considered after the respective primary study is accepted for publication. All data provided are anonymized to respect the privacy of patients who have participated in the trial in line with applicable laws and regulations.

## Notes

### Clinical Trial

NCT04371666

### Author Declarations

The study was conducted in accordance with the Declaration of Helsinki, Good Clinical Practice (GCP), the International Council for Harmonisation E6 Guidance for GCP, and any other applicable local health and regulatory requirements. The study was approved by the respective institutional review boards at each participating study site, and written informed consent was obtained from each patient or their legal guardian. See Table S1 in supplement for full details.

## REFERENCES

[1] Duan D, Goemans N, Takeda S, Mercuri E, Aartsma-Rus A. Duchenne muscular dystrophy. Nat Rev Dis Primers. 2021;7(1):13. doi: 10.1038/s41572-021-00248-3.

[2] Eser G, Topaloglu H. Current outline of exon skipping trials in Duchenne muscular dystrophy. Genes (Basel). 2022;13(7):1241. doi: 10.3390/genes13071241.

[3] Takeda S, Clemens PR, Hoffman EP. Exon-skipping in Duchenne muscular dystrophy. J Neuromuscul Dis. 2021;8(S2):S343–58. doi: 10.3233/JND-210682.

[4] Birnkrant DJ, Bushby K, Bann CM, Apkon SD, Blackwell A, Brumbaugh D, et al. Diagnosis and management of Duchenne muscular dystrophy, part 1: diagnosis, and neuromuscular, rehabilitation, endocrine, and gastrointestinal and nutritional management. Lancet Neurol. 2018;17(3):251–67. doi: 10.1016/S1474-4422(18)30024-3.

[5] Birnkrant DJ, Bushby K, Bann CM, Alman BA, Apkon SD, Blackwell A, et al. Diagnosis and management of Duchenne muscular dystrophy, part 2: respiratory, cardiac, bone health, and orthopaedic management. Lancet Neurol. 2018;17(4):347–61. doi: 10.1016/S1474-4422(18)30025-5.

[6] Broomfield J, Hill M, Chandler F, Crowther MJ, Godfrey J, Guglieri M, et al. Developing a natural history model for Duchenene muscular dystrophy. Pharmacoecon Open. 2024;8(1):79–89. doi: 10.1007/s41669-023-00450-x.

[7] Broomfield J, Hill M, Crowther MJ, Abrams KR. Life expectancy in Duchenne muscular dystrophy: Reproduced individual patient data meta-analysis. Neurology. 2021;97(23):e2304–14. doi: 10.1212/WNL.0000000000012910.

[8] McDonald CM, Henricson EK, Abresch RT, Duong T, Joyce NC, Hu F, et al. Long-term effects of glucocorticoids on function, quality of life, and survival in patients with Duchenne muscular dystrophy: A prospective cohort study. Lancet. 2018;391(10119):451–61. doi: 10.1016/S0140-6736(17)32160-8.

[9] Connolly AM, Florence JM, Zaidman CM, Golumbek PT, Mendell JR, Flanigan MD, et al. Clinical trial readiness in non-ambulatory boys and men with Duchenne muscular dystrophy: MDA-DMD network follow-up. Muscle Nerve. 2016;54(4):681–9. doi: 10.1002/mus.25089.

[10] Gloss D, Moxley RT 3rd, Ashwal S, Oskoui M. Practice guideline update summary: corticosteroid treatment of Duchenne muscular dystrophy: report of the Guideline Development Subcommittee of the American Academy of Neurology. Neurology. 2016;86(5):465–72. doi: 10.1212/WNL.0000000000002337.

[11] Matthews E, Brassington R, Kuntzer T, Jichi F, Manzur AY. Corticosteroids for the treatment of Duchenne muscular dystrophy. Cochrane Database Syst Rev. 2016;2016(5):CD003725. doi: 10.1002/14651858.CD003725.pub4.

[12] Mayer OH, Finkel RS, Rummey C, Benton MJ, Glanzman AM, Flickinger J, et al. Characterization of pulmonary function in Duchenne muscular dystrophy. Pediatr Pulmonol. 2015;50(5):487–94. doi: 10.1002/ppul.23172.

[13] Shiba N, Yang X, Sato M, Kadota S, Suzuki Y, Agata M, et al. Efficacy of exon-skipping therapy for DMD cardiomyopathy with mutations in actin binding domain 1. Mol Ther Nucleic Acids. 2023:102060. doi: 10.1016/j.omtn.2023.102060.

[14] Hoy SM. Delandistrogene moxeparvovec: first approval. Drugs. 2023;83(14):1323–1329.

[15] Lipson KE, Wong C, Teng Y, Spong S. CTGF is a central mediator of tissue remodeling and fibrosis and its inhibition can reverse the process of fibrosis. Fibrogenesis Tissue Repair. 2012;5(Suppl 1):S24. doi: 10.1186/1755-1536-5-S1-S24.

[16] Morales MG, Cabello-Verrugio C, Santander C, Cabrera D, Goldschmeding R, Brandan E. CTGF/CCN-2 over-expression can directly induce features of skeletal muscle dystrophy. J Pathol. 2011;225(4):490–501. doi: 10.1002/path.2952.

[17] Passerini L, Bernasconi P, Baggi F, Confalonieri P, Cozzi F, Cornelio F, et al. Fibrogenic cytokines and extent of fibrosis in muscle of dogs with X-linked golden retriever muscular dystrophy. Neuromuscul Disord. 2002;12(9):828–35. doi: 10.1016/s0960-8966(02)00071-8.

[18] Sun G, Haginoya K, Wu Y, Chiba Y, Nakanishi T, Onuma A, et al. Connective tissue growth factor is overexpressed in muscles of human muscular dystrophy. J Neurol Sci. 2008;267(1-2):48–56. doi: 10.1016/j.jns.2007.09.043.

[19] Connolly AM, Zaidman CM, Brandsema JF, Phan HC, Tian C, Zhang X, et al. Pamrevlumab, a fully human monoclonal antibody targeting connective tissue growth factor, for non-ambulatory patients with Duchenne muscular dystrophy. J Neuromuscul Dis. 2022;10(4):685–99. doi: 10.3233/JND-230019

[20] Wong BL, Mercuri E, Connolly AM, Phan HC, Péréon Y, De Lucio S, et al. Pamrevlumab did not meet its primary endpoint for ambulatory patients with Ducheene Muscular Dystrophy: the LELANTOS-2 trial. 2024. [submitted to J Neuromuscul Dis as companion manuscript].

[21] Seferian AM, Moraux A, Annoussamy M, Canal A, Decostre V, Diebate O, et al. Upper limb strength and function changes during a one-year follow-up in non-ambulant patients with Duchenne Muscular Dystrophy: an observational multicenter trial. PLoS One. 2015;10(2):e0113999. doi: 10.1371/journal.pone.0113999.

