## Supplemental Material for "Pamrevlumab did not meet its primary endpoint for non-ambulatory patients with Duchenne Muscular Dystrophy: the LELANTOS-1 trial"

### LELANTOS-1: SUPPLEMENTARY MATERIAL

#### Methods

A patient was eligible for the study if the following inclusion criteria were met:

I. Age, consent, and contraception:
   a. Males at least 12 years of age who were non-ambulatory at screening initiation
   b. Written consent provided by patient and/or legal guardian
   c. Male patients with partners of childbearing potential had to use contraception during the conduct of the study and for 12 weeks after the last dose of study drug.

II. Duchenne muscular dystrophy (DMD) diagnosis: Medical history included diagnosis of DMD and confirmed Duchenne mutation using a validated genetic test

III. Performance criteria:
   a. Brooke Score for Arms and Shoulders ≤5
   b. Able to undergo magnetic resonance imaging (MRI) test for the upper arm extremities (biceps brachii muscle) and cardiac muscle
   c. Able to perform spirometry

IV. Pulmonary and cardiac criteria:
   a. Average (of Screening and Day 0) percent predicted forced vital capacity (ppFVC) between 45 and 85, inclusive
   b. Left ventricular ejection fraction ≥50% as determined by local cardiac MRI read at screening or within 3 months prior to randomization (Day 0)
   c. If a patient had a history of cardiomyopathy, then he had to be on a stable dose of cardiomyopathy/heart failure medications (e.g., angiotensin converting enzyme inhibitors, aldosterone receptors blockers, angiotensin-receptor blockers, and beta blockers) for at least 1 month prior to screening. If a patient had no diagnosis of cardiomyopathy, then no dose of cardiomyopathy/heart failure medication was required for eligibility.
   d. On a stable dose of systemic corticosteroids for a minimum of 6 months, with no substantial change in dosage for a minimum of 3 months (except for adjustments for changes in body weight) prior to screening. Corticosteroid dosage was in compliance

with the DMD Care Considerations Working Group recommendations (e.g., prednisone or prednisolone 0.75 mg/kg per day or deflazacort 0.9 mg/kg per day) or stable dose [1]. A reasonable expectation was that dosage and dosing regimen would not change significantly for the duration of the study.

V. Vaccination: Agreement to receive annual influenza vaccinations during the study

VI. Laboratory criteria:

a. Adequate renal function: cystatin C ≤1.4 mg/L

b. Adequate hematology and electrolytes parameters:

i. Platelets >100,000/μL

ii. Hemoglobin >12 g/dL

iii. Absolute neutrophil count >1500/μL

iv. Serum calcium (Ca), potassium (K), sodium (Na), magnesium (Mg), and phosphorus (P) levels were within a clinically accepted range for DMD patients.

c. Adequate hepatic function:

i. No history or evidence of liver disease

ii. Gamma glutamyl transferase ≤3× upper limit of normal (ULN)

iii. Total bilirubin ≤1.5×ULN

Patients were excluded if any of the following criteria were met:

I. General criteria:

a. Previous exposure to pamrevlumab

b. Body mass index (BMI) ≥40 kg/$m^2$ or weight >117 kg

c. History of:

i. Allergic or anaphylactic reaction to human, humanized, chimeric, or murine monoclonal antibodies

ii. Hypersensitivity to study drug or any component of study drug

iii. Hypersensitivity reaction to gadolinium-based contrast agents required for MRI acquisition

d. Exposure to any investigational drug (for DMD or not) in the 30 days prior to screening initiation or use of approved DMD therapies (e.g., eteplirsen [exondys

51], ataluren, golodirsen [vyondys 53], casimersen [amondys 45]) within 5 half-lives of screening, whichever was longer, with the exception of systemic corticosteroids (including deflazacort)

II. Cardiac, renal, and pulmonary assessments:

a. Severe uncontrolled heart failure (NYHA Classes III-IV) or renal dysfunction, including any of the following:

i. Need for intravenous diuretics or inotropic support within 8 weeks prior to screening

ii. Hospitalization for a heart failure exacerbation or arrhythmia within 8 weeks prior to screening

iii. Patients with glomerular filtration rate of <30 mL/min/1.73m$^2$ or with other evidence of acute kidney injury as determined by investigator

b. Arrhythmia requiring antiarrhythmic therapy

c. Required ≥16 hours continuous ventilation

d. Hospitalization due to respiratory failure within the 8 weeks prior to screening

e. Poorly controlled asthma or underlying lung disease, such as bronchitis, bronchiectasis, emphysema, or recurrent pneumonia that in the opinion of the investigator might have impacted respiratory function.

III. Clinical judgments:

a. The investigator judged that the patient would be unable to fully participate in the study and complete the study for any reason, including inability to comply with study procedures and treatment, or any other relevant medical or psychiatric conditions that could confound efficacy assessment and/or safety assessment

**Table S1. Institutional review board and ethics approvals for LELANTOS-1**

| Sponsor | Protocol # | Country Name | Recipient Type | Recipient Name | Address | Site: Site # | Ethics approval? (granted/waived) |
|---|---|---|---|---|---|---|---|
| FibroGen | FGCL-3019-093 | AUSTRALIA | Central EC | Research Ethics & Governance RCH Human Research Ethics Committee The Royal Children's Hospital Melbourne | Level 4, South Building 50 Flemington Road Parkville Victoria 3052 Australia | N/A | Granted |
| FibroGen | FGCL-3019-093 | AUSTRIA | Central EC | Ethikkommission der Stadt Wien | 3 Thomas-Klestil-Platz 8/2 1030 Wien Austria | N/A | Granted |
| FibroGen | FGCL-3019-093 | BELGIUM | Local IEC | Commissie voor Medische Ethiek Universiteit Gent/UZ Gent | Commissie voor Medische Ethiek Universiteit Gent/UZ Gent C. Heymanslaan 10 9000 Gent | 8301 | Granted |
| FibroGen | FGCL-3019-093 | BELGIUM | Local IEC | Centre Hospitalier Régional de la Citadelle | Centre Hospitalier Régional de la Citadelle Boulevard du 12ème de Ligne 1 4000 Liège | 8303 | Granted |

| | | | | | | | |
|---|---|---|---|---|---|---|---|
| FibroGen | FGCL-3019-093 | BELGIUM | Central EC | Ethische Commissie Onderzoek UZ/KU Leuven UZ Leuven | Campus Gasthuisberg Herestraat 49 3000 Leuven Belgium | N/A | Granted |
| FibroGen | FGCL-3019-093 | CANADA | Local IRB/IEC | London Health Sciences Centre | 800 Commissioners Rd E, London, ON N6A 5W9, Canada | 8204 | Granted |
| FibroGen | FGCL-3019-093 | CHINA | Local IEC | Drug Clinical Trial Ethics Committee of Peking Union Medical College Hospital, Chinese Academy of Medical Sciences | Conversion Building, Level 7, Research Ward 2, Peking Union Medical College Hospital, No. 1, Shuaifuyuan, Dongcheng District, Beijing, China 100730 | 8601 | Granted |
| FibroGen | FGCL-3019-093 | CHINA | Local IEC | Clinical Trial Ethics Committee of West China Second University Hospital, Sichuan University | No.16 Linjiang Middle Road, Wuhou District, Chengdu, Sichuan, China 610041 | 8603 | Granted |
| FibroGen | FGCL-3019-093 | CHINA | Local IEC | Institutional Review Board of Children's Hospital of Chongqing Medical University | Department of Neurology, 9th Floor, Inpatient Department, No. 20 Jinyu Avenue, Liangjiang New District, Chongqing, | 8604 | Granted |

| | | | | | | | |
|---|---|---|---|---|---|---|---|
| | | | | | Chingqing, China 400015 | | |
| FibroGen | FGCL-3019-093 | CZECH REPUBLIC | Local IEC | Etická komise Fakultní Nemocnice v Motole | Etická komise Fakultní nemocnice v Motole V Úvalu 84 150 06 Praha 5 Czech Republic | 8312 | Granted |
| FibroGen | FGCL-3019-093 | CZECH REPUBLIC | Central EC | Etická komise FN Brno | Jihlavská 20 625 00 Brno Czech Republic | N/A | Granted |
| FibroGen | FGCL-3019-093 | FRANCE | Central EC | Comité de protection des personnes Sud-Méditerranée I | Hôpital Sainte Marguerite Pavillon 9 - 270, bld Sainte Marguerite 13009 MARSEILLE France | N/A | Granted |
| FibroGen | FGCL-3019-093 | ISRAEL | Local IEC | Sheba Medical Center Hospital - Tel Hashomer | Sackler School of Medicine TEL-HASHOMER 5265601, ISRAEL | 8411 | Granted |
| FibroGen | FGCL-3019-093 | ISRAEL | Local IEC | The Edith Wolfson Medical Center | 62 HaLohamim Street Holon, Tel Aviv Postal code 5822012, ISRAEL | 8412 | Granted |
| FibroGen | FGCL-3019-093 | ITALY | Local IEC | Comitato Etico IRCCS Ospedale San Raffaele | Via Olgettina 60, 20132, Milano (MI), Italy | 8222 | Granted |

| | | | | | | | |
|---|---|---|---|---|---|---|---|
| FibroGen | FGCL-3019-093 | ITALY | Local IEC | Comitato Etico dell'IRCCS Ospedale Pedi-atrico Bambino Gesù | Viale di Villa Pamphili 84-100, 00155, Roma (RM), Italy | 8224 | Granted |
| FibroGen | FGCL-3019-093 | ITALY | Local IEC | Comitato Etico IRCCS E.Medea - Sez.Scientifica Associazione La Nostra Famiglia | Via Don Luigi Monza, 20, 23842, Bosisio Parini (LC), Italy | 8225 | Granted |
| FibroGen | FGCL-3019-093 | ITALY | Central EC | Comitato Etico Territoriale (CET) - Comitato Etico Lazio Area 3 c/o Segreteria Tecnico Scientifica all'Ex Collegio Ioanneum | primo piano stanza 221 Largo F. Vito 1 00168 Roma Italy | N/A | Granted |
| FibroGen | FGCL-3019-093 | NETHERLANDS | Central EC | Commissie Mensgebonden Onderzoek regio Arnhem-Nijmegen | p/a Radboudumc, Huispost 628, PO box 9101, 6500 HB Nijmegen The Netherlands | N/A | Granted |
| FibroGen | FGCL-3019-093 | SPAIN | Central EC | Hospital Clínico San Carlos Comité de Ética de la Investigación con Medicamentos | C/Profesor Martín Lagos, s/n. - Puerta G - 4ª Norte Madrid 28040 Madrid Spain | N/A | Granted |

| | | | | | | | |
|---|---|---|---|---|---|---|---|
| FibroGen | FGCL-3019-093 | SWITZERLAND | Central EC | Kantonale Ethikkommission Bern (KEK) | Hörsaaltrakt Pathologie, Eingang 43A, Büro H372 Murtenstrasse 31 3010 Bern Switzerland | N/A | Granted |
| FibroGen | FGCL-3019-093 | UNITED KINGDOM | Central EC | Yorkshire & The Humber - Leeds East Research Ethics Committee | NHSBT Newcastle Blood Donor Centre Holland Drive Newcastle upon Tyne NE2 4NQ United Kingdom | N/A | Granted |
| FibroGen | FGCL-3019-093 | UNITED STATES | Local IRB/IEC | Cincinnati Children's Institutional Review Board | 3333 Burnet Avenue Cincinnati, Ohio, 45229 | 7901 | Granted |
| FibroGen | FGCL-3019-093 | UNITED STATES | Local IRB/IEC | Cincinnati Children's Hospital Medical Center | 3333 Burnet Avenue Cincinnati, Ohio, 45229 | 7901 | Granted |
| FibroGen | FGCL-3019-093 | UNITED STATES | Local IRB/IEC | University of Colorado School of Medicine | 2921 Stockton Blvd Suite 1400 Room 1429 Sacramento, CA 95817 | 7905 | Granted |
| FibroGen | FGCL-3019-093 | UNITED STATES | Local IRB/IEC | Children's Hospital of Philadelphia | 34th Street and Civic Center Boulevard Philadelphia, Pennsylvania, | 7908 | Granted |

| | | | | | | | |
|---|---|---|---|---|---|---|---|
| | | | | | 19104 | | |
| FibroGen | FGCL-3019-093 | UNITED STATES | Local IRB/IEC | Children's Hospital of Philadelphia Institutional Review Board | 3401 Civic Center Boulevard Philadelphia, Pennsylvania, 19104 | 7908 | Granted |
| FibroGen | FGCL-3019-093 | UNITED STATES | Local IRB/IEC | UCLA Office of Human Research Protection Program | 10889 Wilshire Boulevard Los Angeles, California, 90024 | 7913 | Granted |
| FibroGen | FGCL-3019-093 | UNITED STATES | Local IRB/IEC | UT Southwestern William P. Clements Jr. University Hospital | 6201 Harry Hines Boulevard Dallas, Texas, 75235 | 7920 | Granted |
| FibroGen | FGCL-3019-093 | UNITED STATES | Local IRB/IEC | UMass Chan Medical School | 55 Lake Avenue North Worcester, Massachusetts, 01655 | 7951 | Granted |
| FibroGen | FGCL-3019-093 | UNITED STATES | Local IRB/IEC | UC Davis Office of Research | 2921 Stockton Blvd Suite 1400 Room 1429 Sacramento, CA 95817 | 7952 | Granted |
| FibroGen | FGCL-3019-093 | UNITED STATES | Local IRB/IEC | University of California Davis | 2921 Stockton Blvd Suite 1400 Room 1429 Sacramento, CA 95817 | 7952 | Granted |

| | | | | | | | |
|---|---|---|---|---|---|---|---|
| FibroGen | FGCL-3019-093 | UNITED STATES | Local IRB/IEC | University of California Davis Children's Hospital | 2921 Stockton Blvd Suite 1400 Room 1429 Sacramento, CA 95817 | 7952 | Granted |
| FibroGen | FGCL-3019-093 | UNITED STATES | Local IRB/IEC | University of Kansas Medical Center Research Institute IRB in PSO: WCG IRB | 1019 39th Ave SE Suite 120 Puyallup, WA 98374 | 7954 | Granted |
| FibroGen | FGCL-3019-093 | UNITED STATES | Local IRB/IEC | Arkansas Children's Hospital | 1 Children's Way Little Rock, Arkansas, 72202 | 7956 | Granted |
| FibroGen | FGCL-3019-093 | UNITED STATES | Local IRB/IEC | Johns Hopkins Medicine - Office of Human Subjects Research | 1620 McElderry Street East Baltimore Campus Baltimore, Maryland, 21205-1911 | 7959 | Granted |
| FibroGen | FGCL-3019-093 | UNITED STATES | Local IRB/IEC | Kennedy Krieger Institute | 707 North Broadway Baltimore, Maryland, 21205 | 7959 | Granted |
| FibroGen | FGCL-3019-093 | UNITED STATES | Local IRB/IEC | The Johns Hopkins Institute For Clinical And Translational Research | 1620 McElderry Street Reed Hall - BI30 Baltimore, Maryland, 21205-1911 | 7959 | Granted |
| FibroGen | FGCL-3019-093 | UNITED STATES | Local IRB/IEC | Ann & Robert H. Lurie Children's Hospital of Chicago | 225 East Chicago Avenue Chicago, Illinois, 60611 | 7960 | Granted |
| FibroGen | FGCL-3019-093 | UNITED STATES | Local IRB/IEC | Ann and Robert H. Lurie Children's Hospital of Chicago Research | 225 East Chicago Avenue Chicago, Illinois, 60611 | 7960 | Granted |

| | | | | | | | |
|---|---|---|---|---|---|---|---|
| | | | | Center | | | |
| FibroGen | FGCL-3019-093 | UNITED STATES | Local IRB/IEC | University of Utah Institutional Review Board | The Research Administration Building 75 South 2000 East Salt Lake City, Utah 84112 | 7962 | Granted |
| FibroGen | FGCL-3019-093 | UNITED STATES | Local IRB/IEC | Penn State Health Children's Hospital IRB on 1572: WCG IRB | 1019 39th Ave SE Suite 120 Puyallup, WA 98374 | 7965 | Granted |
| FibroGen | FGCL-3019-093 | UNITED STATES | Local IRB/IEC | University of Michigan IRB on 1572: WCG IRB | 1019 39th Ave SE Suite 120 Puyallup, WA 98374 | 7966 | Granted |
| FibroGen | FGCL-3019-093 | UNITED STATES | Local IRB/IEC | Vanderbilt University Medical Center | Human Research Protections Program 3319 West End Ave Suite 600 Nashville, TN 37203 | 7968 | Granted |
| FibroGen | FGCL-3019-093 | UNITED STATES | Local IRB/IEC | University of Iowa Institutional Review Board/Human Subjects Office IRB in PSO/1572: WCG IRB | 1019 39th Ave SE Suite 120 Puyallup, WA 98374 | 7969 | Granted |
| FibroGen | FGCL-3019-093 | UNITED STATES | Local IRB/IEC | Corewell Health (Spectrum Health) Hospitals Helen DeVos Children's | 35 Michigan Street Northeast Mail Code 161 Grand Rapids, Michigan, 49503 | 7971 | Granted |

| | | | | | | | |
|---|---|---|---|---|---|---|---|
| | | | | Hospital | | | |
| FibroGen | FGCL-3019-093 | UNITED STATES | Local IRB/IEC | Spectrum Health Institutional Review Board | 15 Michigan Street NE Suite 701 Grand Rapids, MI 49503 | 7971 | Granted |
| FibroGen | FGCL-3019-093 | UNITED STATES | Central IRB/IEC | Advarra | 6940 Columbia Gateway Drive Columbia, Maryland, 21046 | 7958 | Granted |
| FibroGen | FGCL-3019-093 | UNITED STATES | Central IRB/IEC | WCG IRB | 1019 39th Ave SE Suite 120 Puyallup, WA 98374 | N/A | Granted |
| FibroGen | FGCL-3019-093 | UNITED STATES | Central IRB/IEC | WCG IRB - Copernicus Group Independent Review Board | 1019 39th Ave SE Suite 120 Puyallup, WA 98374 | 7970 | Granted |
| FibroGen | FGCL-3019-093 | UNITED STATES | Central IRB/IEC | WCG IRB - Western Institutional Review Board | 1019 39th Ave SE Suite 120 Puyallup, WA 98374 | N/A | Granted |
| FibroGen | FGCL-3019-093 | UNITED STATES | Central IRB/IEC | Western Institutional Review Board - Local IRB | 1019 39th Avenue Southeast Suite 120 Puyallup, Washington, 98374-2215 | 7920 | Granted |
| FibroGen | FGCL-3019-093 | UNITED STATES | Central IRB/IEC | WIRB-Copernicus Group - WCG Global Headquarters - North America | 1019 39th Avenue Southeast Suit 120 Puyallup, Washington, 98374-2215 | N/A | Granted |

### REFERENCE


1. Gloss D, Moxley RT 3rd, Ashwal S, Oskoui M. Practice guideline update summary: corticosteroid treatment of Duchenne muscular dystrophy: Report of the Guideline Development Subcommittee of the American Academy of Neurology. Neurology. 2016;86(5):465–72. doi:10.1212/WNL.0000000000002337
